# High-sensitivity detection of *Mycobacterium tuberculosis* DNA in tongue swab samples

**DOI:** 10.1101/2024.07.26.24311064

**Authors:** Alaina M. Olson, Rachel C. Wood, Kris M. Weigel, Alexander J. Yan, Katherine A. Lochner, Rane B. Dragovich, Angelique K. Luabeya, Paul Yager, Mark Hatherill, Gerard A. Cangelosi

**Author notes:** Corresponding author (GC).

## Abstract

Tongue swab (TS) sampling combined with qPCR to detect *Mycobacterium tuberculosis* (MTB) DNA is a promising alternative to sputum testing for tuberculosis (TB) diagnosis. In prior studies, the sensitivity of tongue swabbing has usually been lower than sputum. In this study, we evaluated two strategies to improve sensitivity. In one, centrifugation was used to concentrate tongue dorsum bacteria from 2-mL suspensions eluted from high-capacity foam swab samples. The pellets were resuspended as 500-µL suspensions, and then mechanically lysed prior to dual-target qPCR to detect MTB insertion elements IS*6110* and IS*1081*. Fractionation experiments demonstrated that most of the MTB DNA signal in clinical swab samples (99.22% ± 1.46%) was present in the sedimentable fraction. When applied to archived foam swabs collected from 124 South Africans with presumptive TB, this strategy exhibited 83% sensitivity (71/86) and 100% specificity (38/38) relative to sputum MRS (microbiological reference standard; sputum culture and/or Xpert® Ultra). The second strategy used sequence-specific magnetic capture (SSMaC) to concentrate DNA released from MTB cells. This protocol was evaluated on archived Copan FLOQSwabs® flocked swab samples collected from 128 South African participants with presumptive TB. Material eluted into 500 µL buffer was mechanically lysed. The suspensions were digested by proteinase K, hybridized to biotinylated dual-target oligonucleotide probes, and then concentrated ∼20-fold using magnetic separation. Upon dual-target qPCR testing of concentrates, this strategy exhibited 90% sensitivity (83/92) and 97% specificity (35/36) relative to sputum MRS. These results point the way toward automatable, high-sensitivity methods for detecting MTB DNA in TS.

**Importance:** Improved testing for tuberculosis (TB) is needed. Using a more accessible sample type than sputum may enable the detection of more cases, but it is critical that alternative samples be tested appropriately. Here, we describe two new, highly accurate methods for testing tongue swabs for TB DNA.

## Introduction

Screening and diagnosis of pulmonary tuberculosis (TB) remain urgent global health priorities. The standard patient sample for microbiological diagnosis of TB is sputum, a viscous expectorated material. The availability of alternatives to sputum would greatly facilitate testing for active TB in both clinical and community settings (1–4). One such alternative is tongue swab (TS) testing (5–12). In TS testing, the dorsum of the tongue is swabbed and the collected material is tested by qPCR for MTB DNA. Tongue swabbing is fast, painless, and does not require facilities for privacy or aerosol control. In a recent study of user preference, most health care workers expressed their preference for swabbing over sputum collection (13). Individuals who may struggle to produce sputum, such as children and people living with HIV (PLHIV), can easily provide swabs in any setting.

TS testing has exhibited varying sensitivities relative to sputum microbiological reference standards (sputum MRS), ranging from ∼75% when using commercial sputum testing platforms (5, 6), to >90% when swabs are tested by manual, laboratory-based methods designed specifically for swab testing (5, 14). Specificity relative to sputum MRS consistently exceeds 95% (5, 6, 14). Recently, Steadman et al. (14) reported a laboratory-based method with high sensitivity. Their protocol incorporated heat, high-energy bead-beating with a BioSpec instrument, and high-volume qPCR targeting MTB insertion elements IS*6110* and IS*1081*.

Although costly and potentially difficult to automate, this approach yielded high accuracy (93% sensitivity and 99% specificity relative to sputum MRS) when applied to symptomatic participants in a clinical setting in Uganda (14). Such observations illustrate the biological potential for TS as a non-sputum sample when appropriate analytical methods are used.

The current study evaluated two strategies to maximize TS sensitivity, using methods designed to be feasible to automate. The first strategy aimed to collect a greater proportion of the available MTB bacilli. Previous work suggested that only a fraction of available tongue dorsum biomass was collected with nylon flocked swabs, and that a substantial portion is left behind (8). In the current study, tongue dorsum samples were collected with high-capacity open-cell foam swabs instead. Preliminary experiments showed that Medline Pint-Size™ DenTips® Oral Swabs collect more tongue dorsum biomass (measured as absorbance at 260 nm) than Copan FLOQSwabs® flocked swabs (p < 0.0001; Supplemental material S1). We conjectured that this would enable the collection of more MTB bacilli from tongue dorsa.

However, increasing the size of the swab head presents a new challenge. Compared to the 0.5 mL buffer volume used with FLOQSwabs, a larger buffer volume of 2 mL is needed to release collected biomass into the foam swab sample eluate. To mitigate sample dilution, we evaluated cell concentration by centrifugation prior to cell disruption. Concentrated suspensions were mechanically lysed, and the crude lysate was amplified by a high-volume qPCR.

In this study, we applied this method to archived foam swab samples that were previously collected from a cohort of South African participants with presumptive TB (N = 124).

Replicate FLOQSwabs from the same participants were previously tested on two commercial sputum testing platforms: Cepheid Xpert® MTB/RIF Ultra (Xpert Ultra) and Molbio Truenat® MTB Ultima (MTB Ultima). Xpert Ultra was 75.5% sensitive and 100% specific, and MTB Ultima was 71.6% sensitive and 96.9% specific, relative to sputum MRS (5). We investigated whether sensitivity was improved by the alternate sample collection and processing method using foam swabs. In the process, we also investigated whether the MTB DNA signal present in clinical swab samples resides in the sedimentable (particulate) fraction, rather than in the soluble (supernatant) fraction.

In the second strategy, we focused on extracting and concentrating DNA after release from bacterial cells, rather than concentrating bacterial cells. After collection, tongue dorsum biomass on a FLOQSwab head is typically eluted into ∼500 µL of TE buffer (5–9, 14). Because a FLOQSwab collects just ∼80 µL of oral material, it is critical that all, or nearly all, of this material be included in the qPCR reaction. In past studies we have used ethanol precipitation of DNA for this purpose (8, 9). However, this method is cumbersome and difficult to automate. Recently, Oreskovic et al. (15) reported the use of hybridization probes immobilized on magnetic beads to selectively capture and concentrate MTB DNA from dilute urine samples. This strategy, which we term sequence-specific magnetic capture (SSMaC), has several potential advantages. First, it is facile and amenable to automation in POC devices (16). For example, the fully automated, POC DASH™ (Digital Analyzer for Selective Hybridization) Rapid PCR system (Nuclein, Inc., Austin, TX, USA) incorporates a 10-min SSMaC step that concentrates SARS-CoV-2 RNA in swab samples, enabling qPCR detection with high sensitivity (17). Recently, a lab-based MTB-targeted version of the DASH assay was described for sputum testing (18). Second, in addition to concentrating target DNA, SSMaC enables partial purification of target DNA by removing qPCR inhibitors.

To evaluate the effectiveness of SSMaC for tongue swab testing, we optimized a previously reported (19) SSMaC protocol and applied it to archived clinical FLOQSwab samples that were collected from a cohort of South African participants with presumptive TB (N = 128).

Replicate FLOQSwab samples from these participants were previously evaluated by Xpert Ultra and Molbio Ultima (5). The current study investigated whether samples from the same participants exhibited better sensitivity when analyzed by the optimized SSMaC-based method.

## Materials and methods

### Ethics

This study was approved by the University of Cape Town Human Research Ethics Committee (reference number 160/202) and the University of Washington Human Subjects Division (STUDY00001840).

### Study setting and participants

Enrollment and sample collection were described previously (5). Enrollment began July 2021 and continued until March 2023 in Worcester, South Africa.

### Foam swab collection

Participants with presumptive TB were enrolled in two cohorts in Worcester, South Africa, as described (5). When Cohort 2 (5) was enrolled, a subset (N = 156) provided foam swab samples (Pint-Size™ DenTips® Oral Swabs, MDS096202P; Medline Industries LP, Northfield, IL, USA) in addition to flocked swabs (regular FLOQSwabs® Flocked Swabs, 520CS01; Copan Italia, Brescia, IT). Foam swabs from some of these participants (N = 31) were used in unrelated studies, and one (N = 1) was lost during processing due to a broken collection tube. All remaining foam swabs (N = 124) were used in this study (Figure 1).

**Figure 1.**
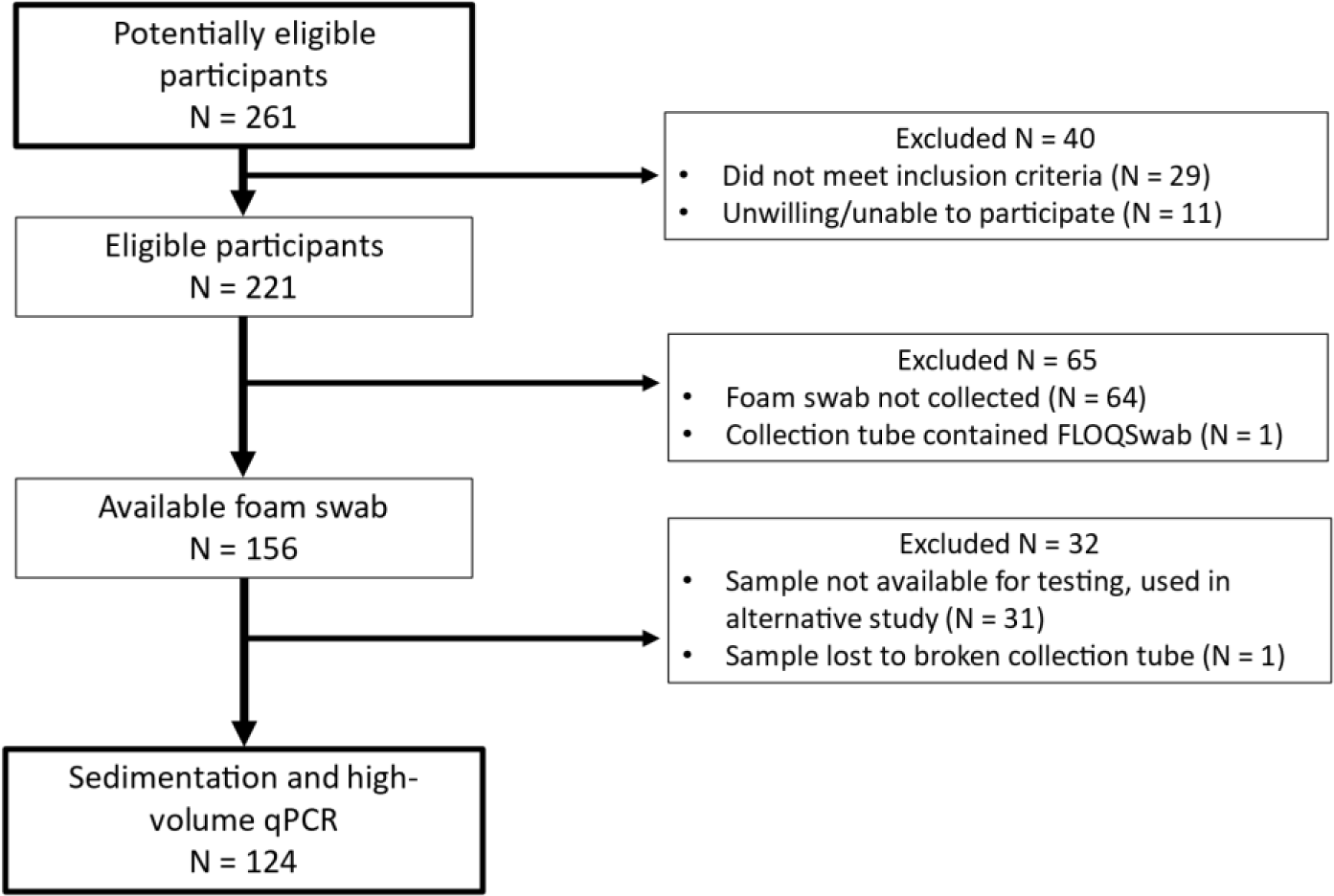
Foam swab STARD Diagram

All swab samples were collected prior to sputum collection. Sample collection occurred over two days, with high-capacity foam swabs collected after two FLOQSwabs on Day 2. Foam swabs were brushed against the dorsum of the tongue for 15 sec and stored dry in 5 mL tubes (6101S, Globe Scientific Inc, Mahwah, NJ, USA). Swab shafts were cut with scissors to fit into the tubes. Samples were stored at -80 °C and shipped on dry ice to Seattle, Washington for testing at the end of the study.

### Foam swab processing

Foam swabs were kept at -80 °C until ready for processing. Swabs were removed from the freezer and immediately boiled at 100 °C in a water-filled heat block (88-860-023; Thermo Fisher Scientific Inc., Waltham, MA, USA) for 10 min to inactivate pathogens. Batches were limited to 4 tubes per block (88-860-107; Thermo Fisher Scientific Inc., Waltham, MA, USA), for a total of 8 at a time, to prevent excess temperature drops. 2 mL TE buffer (pH 8.0) was added to the sample followed by a 30 sec vortex at max speed (Vortex-Genie 2, SI-0236; Scientific Industries, Inc, Bohemia, NY, USA). During sample aspiration using a p1000 micropipette tip, the tip was used to compress the foam head against the wall of the tube to maximize liquid recovery. All of the recoverable liquid sample was transferred to a 2-mL screw-cap tube (02707355; Thermo Fisher Scientific Inc., Waltham, MA, USA). Samples were centrifuged at 17,320 RCF at room temperature for 2 min. After sedimentation, 1.5 mL supernatant was removed from the uppermost portion of the sample, avoiding the pellet, and then discarded. Samples were re-spun and aspirated again if the pellet was noticeably disturbed during aspiration. The pellet was resuspended in the remaining ∼500 µL supernatant. Bacilli were then mechanically lysed as follows. Working in batches of 16 (including one positive and negative extraction control swab each), 150 mg pre-weighed 0.1 mm glass disruption beads (9830; Research Products International (RPI), Mt. Prospect, IL, USA) were added to the sample tube containing the suspension. Samples were bead-beaten for three rounds of 1 min on with 1 min rests, using a high-energy cell disruptor (Mini-Beadbeater-16, 607; BioSpec Products, Inc., USA). Samples were spun down briefly. While avoiding the glass beads, the entire recoverable liquid volume was transferred to a 1.5-mL snap-cap tube (05408129; Thermo Fisher Scientific Inc., Waltham, MA, USA). Samples were stored at 4 °C briefly before plating for same-day qPCR.

Extraction controls were prepared using swabs collected from healthy volunteers in Seattle; the positive controls were spiked with 40,000 cells of log-phase cultured *M. tuberculosis* H37Ra (ATCC), in 40 µL PBSGT (1x Phosphate-Buffered Saline, 15% Glycerol, 0.05% Tween- 80). All were stored at –80 °C prior to processing.

### Foam swab testing

Fifty microliters of crude lysate were plated in technical duplicates, each mixed with 10 µL of master mix comprising 6 µL 10x KAPA3G HotStart Master (09084711103; Roche, Basel, CH), 500 nM final concentration of each primer (Integrated DNA Technologies, Coralville, IA, USA), 250 nM final concentration of each probe (Integrated DNA Technologies, Coralville, IA, USA), and 2.5 µL molecular grade water, for a final reaction volume of 60 µL. Thermal cycling was performed using the Bio-Rad CFX96 Deep Well Real-Time PCR Detection System (1854095, Bio-Rad Laboratories, Inc., Hercules, CA, USA) with the following cycling parameters from Steadman et al. (14): Initial denaturation at 98 °C for 3 min, followed by 45 cycles of, 1. Denaturation at 98 °C for 5 sec, 2. Annealing at 64 °C for 20 sec, and 3.

Elongation at 72 °C for 30 sec, concluding with a final elongation at 72 °C for 1 min. All ramp rates were 2.5 °C per sec. Results were exported from the Bio-Rad CFX96 Deep Well machine using a threshold of 1000 RFU to Microsoft Excel for further analysis and calculations. Cq and quantification values were averaged between the two replicates of each sample.

### FLOQSwab collection

Participants were enrolled and sampled as previously described (5) and shown here in a STARD diagram (Figure 2).

**Figure 2.**
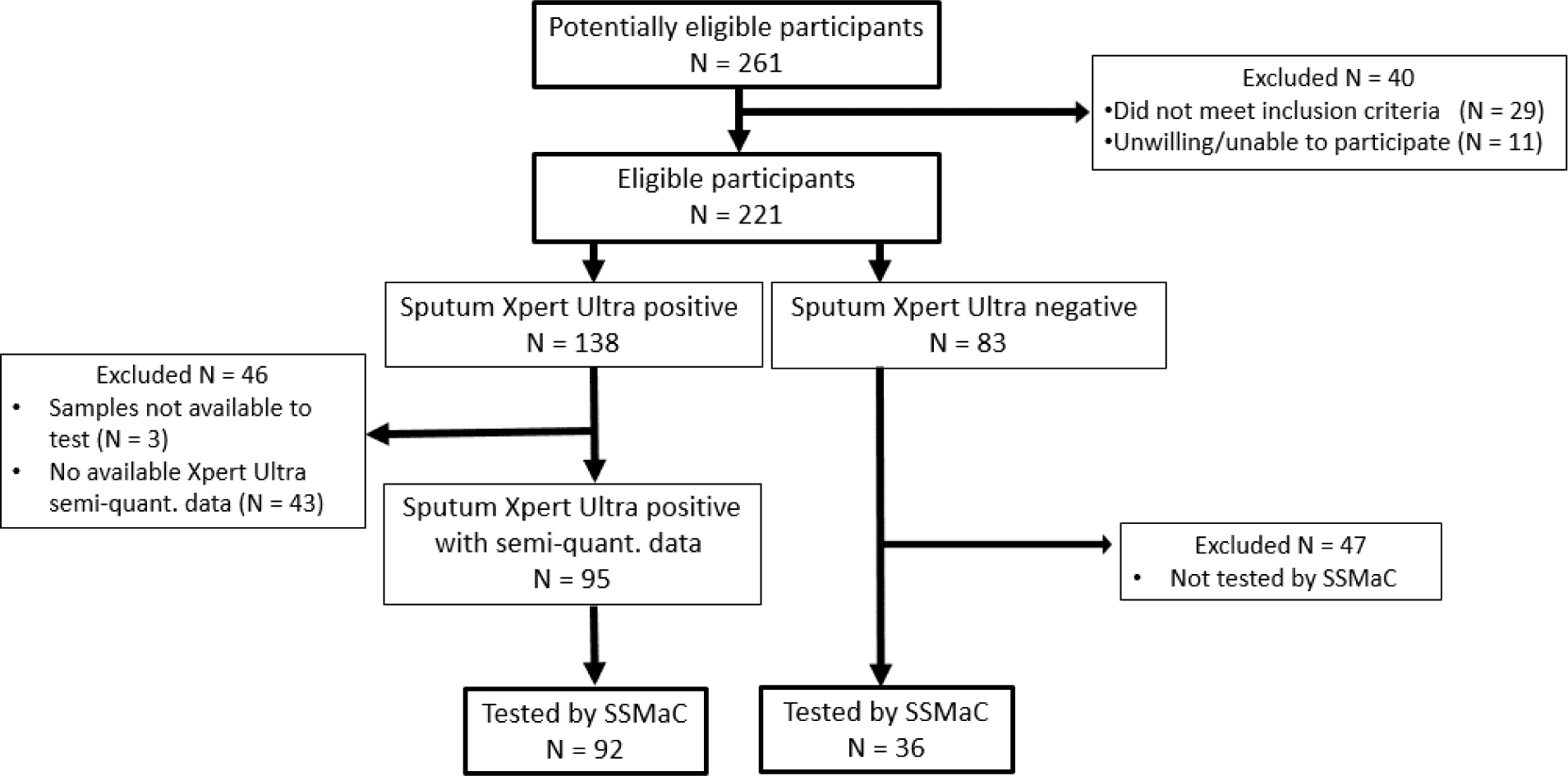
FLOQSwab STARD Diagram

In Cohort 2 of the previous study (5), a subset of participants (N = 92) had available sputum Xpert Ultra (SPUXU) semiquantitative data (trace, very low, low, moderate, high). Samples from the 92 participants with SPUXU data were used for the current study. SPUXU semi-quantitative (SQ) and quantitative Cq results were exported and cleaned by members of the team at the clinical site. Of the 92 participants, 42 were SPUXU high, 16 medium, 23 low, 8 very low and 3 trace. Six of these 92 participants were sputum culture negative.

### FLOQSwab sample processing by SSMaC

Samples for the current study were processed in batches of 10 clinical samples alongside one positive and negative processing control each for a total of 12 samples. The positive controls were swabs collected from healthy donors in Seattle, WA and then spiked with 10 µL of buffer containing 10,000 cells of log-phase cultured *Mycobacterium tuberculosis* H37Ra suspended in PBSGT. The negative controls were swabs collected from healthy donors in the same way and stored without spiking.

Samples previously processed for and tested by Molbio Truenat® MTB Ultima (5) remained in excess (∼494 µL) and were stored at -80 °C. These archived lysates were thawed and ∼474 µL aliquots were subjected to a 30 min proteinase K digestion at 55 °C. The enzyme and any present DNA were then denatured for 10 min at 100 °C. The denatured DNA was hybridized to biotinylated oligonucleotide capture probes for 20 min at 55 °C as previously described (19) with modifications to probe sequences (Table S2). A second set of probes targeting an additional insertion sequence, IS*1018*, was added in equimolarity to an IS*6110* set based on sequences known to work in combination (for qPCR) as shown by Steadman et al. (14) (Supplemental material 1). These sequences align with the sequences used for downstream amplification. Twenty microliters of washed magnetic beads (Dynabeads™ MyOne™ Streptavidin C1, 65001; Thermo Fisher Scientific Inc., Waltham, MA, USA) were used to capture the biotinylated oligonucleotides hybridized with MTB DNA. Magnetic concentration was used to purify and concentrate MTB DNA 20-25-fold, resulting in 20 µL of eluate as previously described (19). This process involved washing away parts of the sample not bound to magnetic beads, including non-target DNA, cell debris, swab flocking, any carried-over disruption beads from lysis, salts, surfactants, and other potential PCR inhibitors. Finally, a 5 min heat-block-incubation at 95 °C released the purified target DNA into 20 µL TE buffer.

### Flocked swab sample qPCR reaction and analysis

Following SSMaC processing, five microliters of concentrated bead eluate (MTB DNA) were amplified in a 20 µL qPCR reaction adapted from Steadman et al. (14) (Supplemental material S2). Changes included modifying the IS*1081* probes with the fluorescein (6-FAM) fluorophore in place of Yakima Yellow®, and excluding RNase P primers and probe. Master mix was plated as 15 µL with the following composition: Final concentrations of 500 nM for each primer and 250 nM for each probe (see table for sequences), 2 µL KAPA3G HotStart Master, 10x concentrated, and 12.5 µL molecular grade water. Each qPCR run included a standard curve of purified MTB DNA and negative controls.

Reactions were run on an Applied Biosystems™ StepOnePlus™ thermal cycler (4376600; Thermo Fisher Scientific Inc., Waltham, MA, USA), with the cycling parameters described above. Normalization included exporting all runs at a 2239 ΔRn threshold. Results were exported from the StepOnePlus™ software to excel, where calculations were executed. Any Cq value below 40 was considered a positive result. For analyses of average Cqs, samples that were undetected were assigned Cq values of 45.

## Results

### Evaluation strategies

The two new methods evaluated were 1) foam swabs combined with sedimentation, and 2) FLOQSwabs processed by SSMaC. Different evaluation strategies were used based on the availability of archived samples. To assess foam swabs with sedimentation, percent sensitivity and specificity relative to sputum MRS were calculated and assessed in relation to values previously reported using other methods (5). For FLOQSwabs processed by SSMaC, we applied the new method to lysates previously processed for Molbio Ultima testing (5). We asked whether sensitivity would be improved by using SSMaC to test the remaining, larger fraction of lysate and compared the results to the original Molbio Ultima results.

### Foam swabs and sedimentation

Foam swabs collected from participants with presumptive TB (N = 124) exhibited 82.6% sensitivity (71/86) and 100% specificity (38/38) relative to sputum MRS (Table 1). These values were nominally higher than previously reported for replicate FLOQSwabs collected from the same 124 participants and tested by Molbio Ultima and Xpert Ultra (5). However, the 95% confidence intervals of all three methods overlap and the differences were not statistically significant by paired z-test (*p* = 0.34 and 0.44, respectively).

**Table 1.** Sensitivities and specificities of methods paired to foam data.

|  | Foam Sedimentation<br>(N = 124) |  | TS Molbio Ultima<br>(N = 121) |  | TS Xpert Ultra<br>(N = 121) |  |
| --- | --- | --- | --- | --- | --- | --- |
| % Sensitivity<br>[95% CI] | 71/86 | 82.6 %<br>[72.87, 89.90] | 66/86 | 76.7%<br>[66.39, 85.18] | 67/86 | 77.9%<br>[67.67, 86.14] |
| % Specificity<br>[95% CI] | 38/38 | 100%<br>[90.75, 100.00] | 34/35 | 97.1%<br>[85.08, 99.93] | 35/35 | 100%<br>[90.00, 100.00] |

SPUXU data were available for a subset (N = 67) of the participants tested by foam swab and sedimentation (Table 2). The method performed best in participants with stronger SPUXU signals. It was insensitive when applied to samples from SPUXU “very low” and “trace” participants, as were the other two methods.

**Table 2.** Sensitivities for each method stratified by sputum Xpert Ultra semi-quantitative (SPUXU SQ) results.

| SPUXU SQ<br>(N) | Foam Sedimentation |  | TS Molbio Ultima |  | TS Xpert Ultra |  |
| --- | --- | --- | --- | --- | --- | --- |
|  | N | % Sensitivity<br>[95% CI] | N | % Sensitivity<br>[95% CI] | N | % Sensitivity [95%<br>CI] |
| High (30) | 30/30 | 100.00%<br>[88.43, 100.00] | 29/30 | 96.67%<br>[82.78, 99.92] | 30/30 | 100.00%<br>[88.43, 100.00] |
| Medium (12) | 12/12 | 100.00%<br>[73.54, 100.00] | 11/12 | 91.67%<br>[61.52, 99.79] | 12/12 | 100.00%<br>[73.54, 100.00] |
| Low (17) | 13/17 | 76.47%<br>[50.10, 93.19] | 11/17 | 64.71%<br>[38.33, 85.79] | 11/17 | 64.71%<br>[38.33, 85.79] |
| Very low (5) | 3/5 | 60.00%<br>[14.66, 94.73] | 3/5 | 60.00%<br>[14.66, 94.73] | 3/5 | 60.00%<br>[14.66, 94.73] |
| Trace (3) | 0/3 | 0.00%<br>[0.00, 70.76] | 0/3 | 0.00%<br>[0.00, 70.76] | 0/3 | 0.00%<br>[0.00, 70.76] |
| **All** | **(58/67)** | **86.57%**<br>**[76.03, 93.67]** | **(54/67)** | **80.60%**<br>**[69.11, 89.24]** | **(56/67)** | **83.58%**<br>**[72.52, 91.51]** |

These results suggest that a significant portion of the MTB DNA signal present in clinical foam swab samples was in the sedimentable fraction, rather than in the supernatants. To assess this quantitatively, we conducted fractionation experiments in which we compared MTB DNA quantities measured by qPCR in concentrated samples relative to the supernatants. In this paired analysis (N = 6 samples), MTB DNA quantities in the pellets were consistently greater than those in the supernatants (p < 0.05, one-sided Wilcoxon Signed Rank test, *W* value = 0). On average, 99.22 ± 1.46% of the available MTB DNA signal was in the sedimentable fraction (Figure 3).

**Figure 3.**
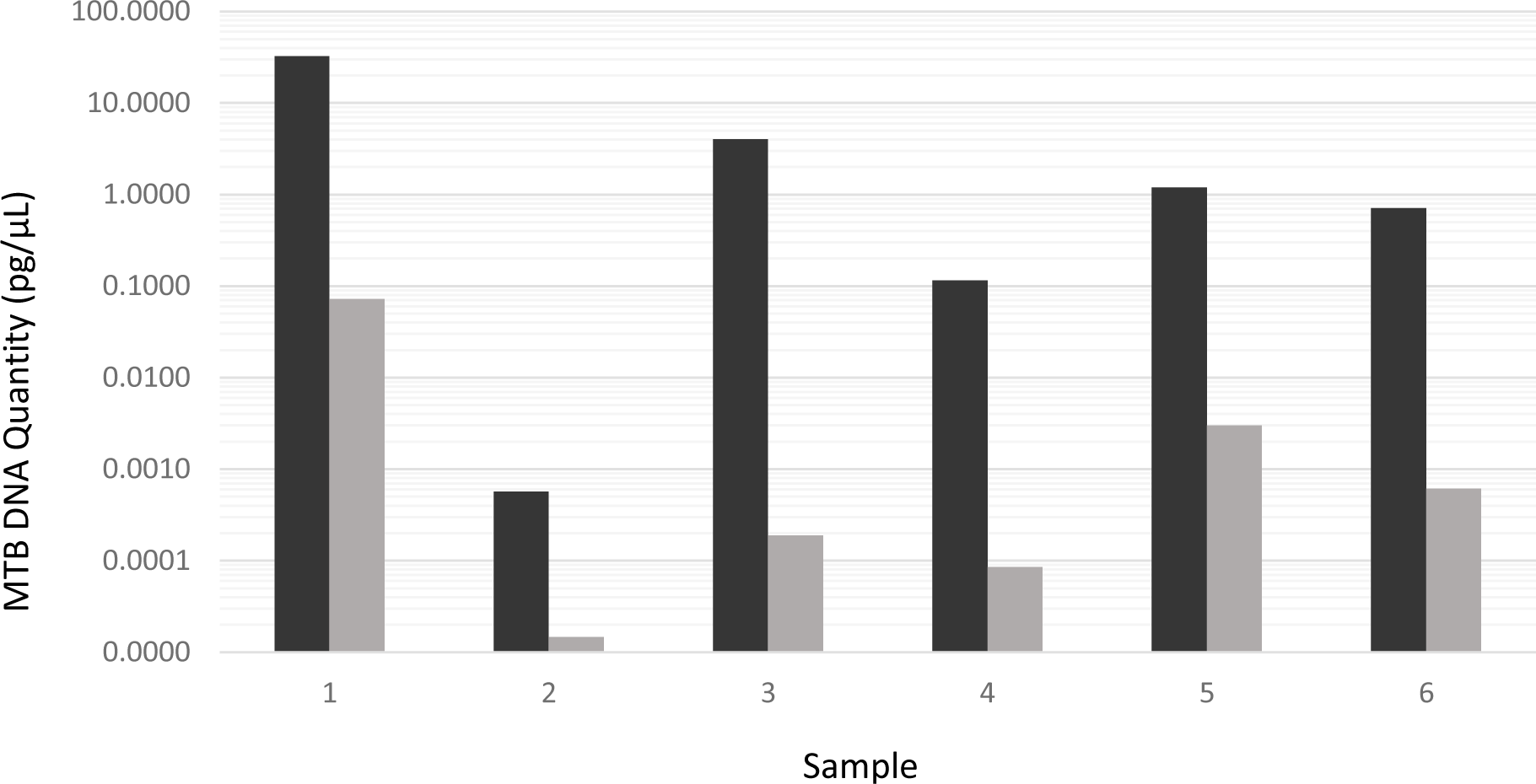
**Paired qPCR results (MTB DNA Quantity pg/µL) for foam swab samples with (black bars) and without (gray bars) sedimentation concentration**

### SSMaC

SSMaC was performed on the lysates from 92 sputum MRS-positive and 36 sputum MRS-negative participants. The sensitivity was 83/92 (90.2%) relative to sputum MRS, a significant improvement over the sensitivity of Molbio Ultima testing (z-test, *p* = 0.04). It was also better than the sensitivity exhibited by Xpert Ultra testing of replicate swabs from these participants, although that difference was not significant (*p* = 0.19) (Table 4, bottom line). The 95% confidence intervals for all three methods overlap with varying severity.

**Table 4.** Percent sensitivity of TS methods relative to SPUXU SQ.

| Sputum Xpert Ultra Semi-Quantity (N) | TS Xpert Ultra |  | TS Molbio Ultima |  | TS SSMaC |  |
| --- | --- | --- | --- | --- | --- | --- |
|  | N | % Sensitivity [95% CI] | N | % Sensitivity [95% CI] | N | % Sensitivity [95% CI] |
| High (42) | 42/42 | 100.00% [91.59, 100.00] | 40/42 | 95.24% [83.84, 99.42] | 42/42 | 100.00% [91.59, 100.00] |
| Medium (16) | 16/16 | 100.00% [79.41, 100.00] | 14/16 | 87.50% [61.65, 98.45] | 15/16 | 93.75% [69.77, 99.84] |
| Low (23) | 16/23 | 69.57% [47.08, 86.79] | 15/23 | 65.22% [42.73, 83.62] | 19/23 | 82.61% [61.22, 95.05] |
| Very Low (8) | 3/8 | 37.50% [8.52, 75.51] | 4/8 | 50.00% [15.70, 84.30] | 6/8 | 75.00% [34.91, 96.81] |
| Trace (3) | 0/3 | 0.00% [0.00, 70.76] | 0/3 | 0.00% [0.00, 70.76] | 1/3 | 33.33% [0.84, 90.57] |
| **All (92)** | **77/92** | **83.70% [74.54, 90.58]** | **73/92** | **79.35% [69.64, 87.08]** | **83/92** | **90.22% [82.24, 95.43]** |

When the results were stratified according to SPUXU SQ, SSMaC consistently trended towards higher sensitivities across all categories of sputum positivity (Table 4). The specificity of SSMaC was also high at 97.22% as compared to TS Molbio Ultima (100%) and TS Xpert Ultra (100%) (Table 5).

**Table 5.** Percent specificity of methods paired by SSMaC.

| TS Xpert Ultra |  | TS Molbio Ultima |  | SSMaC Concentration |  |
| --- | --- | --- | --- | --- | --- |
| N | % Specificity [95% CI] | N | % Specificity [95% CI] | N | % Specificity [95% CI] |
| 36/36 | 100.00% [90.26-100.00] | 36/36 | 100.00% [90.26-100.00] | 35/36 | 97.22% [85.47-99.93] |

### Associations with HIV coinfection status

Sensitivities of both the SSMaC (Table 6) and foam swab (Table 7) methods were higher in participants without HIV coinfections compared to PLHIV. This observation was statistically significant as suggested by two-population z-tests (*p* = 0.04 and 0.0005, respectively). Both methods also had lower average Cqs (stronger MTB DNA signals) among those without HIV. One-tailed, unpaired t-tests suggested significant differences (*p* < 0.0001 and *p* = 0.0061, respectively).

**Table 6.** SSMaC % sensitivity by HIV Status.

| HIV Status | N | % Sensitivity [95% CI] | Two population z-test p value | Cq Avg ± Stdev | One-tailed, unpaired t-test p value |
| --- | --- | --- | --- | --- | --- |
| Positive | 24/30 | 80.00% [61.43, 92.29] | 0.0434 | 36.32 ± 8.08 | <0.0001 |
| Negative | 60/64 | 93.75% [84.76, 98.27] |  | 30.20 ± 5.64 |  |

**Table 7.** Foam swab % sensitivity by HIV Status.

| HIV Status | N | % Sensitivity [95% CI] | Two population z-test *p* value | Cq Avg ± Stdev | One-tailed, unpaired t-test *p* value |
| --- | --- | --- | --- | --- | --- |
| Positive | 16/27 | 59.26% [38.80, 77.61] | 0.0005 | 32.05 ± 7.07 | 0.0061 |
| Negative | 55/59 | 93.22% [83.54, 98.12] |  | 28.06 ± 6.72 |  |

## Discussion

Many exploratory evaluations of TS have used in-house manual nucleic acid amplification tests (NAATs), primarily qPCR, to detect MTB DNA in tongue swab samples (7–9, 14, 20–22). Some of these methods exhibited >90% sensitivity and specificity relative to sputum MRS (8, 9, 14). These observations illustrate the potentially high accuracy of TS testing relative to sputum testing. Other evaluations used the commercial Cepheid Xpert MTB/RIF® Ultra automated qPCR system (Xpert Ultra) (6, 23–26). These approaches generally exhibited lower sensitivity, possibly because the Xpert Ultra is engineered to test sputum rather than swabs.

There remains a need for new methods that are 1) highly sensitive; and 2) amenable to automation for future POC testing. This study evaluated two such methods.

One method used a high-capacity foam swab that collects more material from the tongue dorsum than a Copan FLOQSwab, based on observations by spectrophotometry. Using *A*260 as a measure of biomass picked up from the tongue, foam swabs outperformed FLOQSwabs in a preliminary experiment (Supplemental material S1). We hypothesized that the higher-capacity foam swabs may also collect more TB bacilli from the tongue. To process the foam swabs, we employed a sedimentation step to concentrate the TB cells in a pellet prior to lysis. The sensitivity of the foam swabs was higher than those of the FLOQSwabs when tested by either the Molbio Ultima or Xpert Ultra.

We employed an updated SSMaC protocol in the second method to explore whether further processing of residual crude lysates would detect a greater number of TB positive participants. We concentrated the remaining lysate from FLOQSwabs previously evaluated by Molbio Ultima (5) using optimized SSMaC and tested them by manual qPCR. The resulting sensitivity matched or outperformed those previously reported (5). As reported previously using other methods (9, 20), both new methods exhibited higher sensitivity in HIV-uninfected people.

Figure 4 shows the assay input fraction for the different TS tests evaluated in this study. Using the method described previously (5), Molbio Ultima tested only 1.2% of the total sample volume (Figure 4C). In contrast, SSMaC and foam swab sedimentation tested larger fractions (20% and 25%, respectively). This could help to explain the improvement in sensitivity observed in the latter two methods. Both new methods also appeared to be more sensitive than Xpert Ultra testing of oral swabs. This sensitivity enhancement cannot be attributed to sample volume, because Xpert Ultra tests 100% of the sample (Figure 4A), but may be explained by the limitations of using a commercial platform that is optimized to analyze sputum samples to test swab samples.

**Figure 4.**
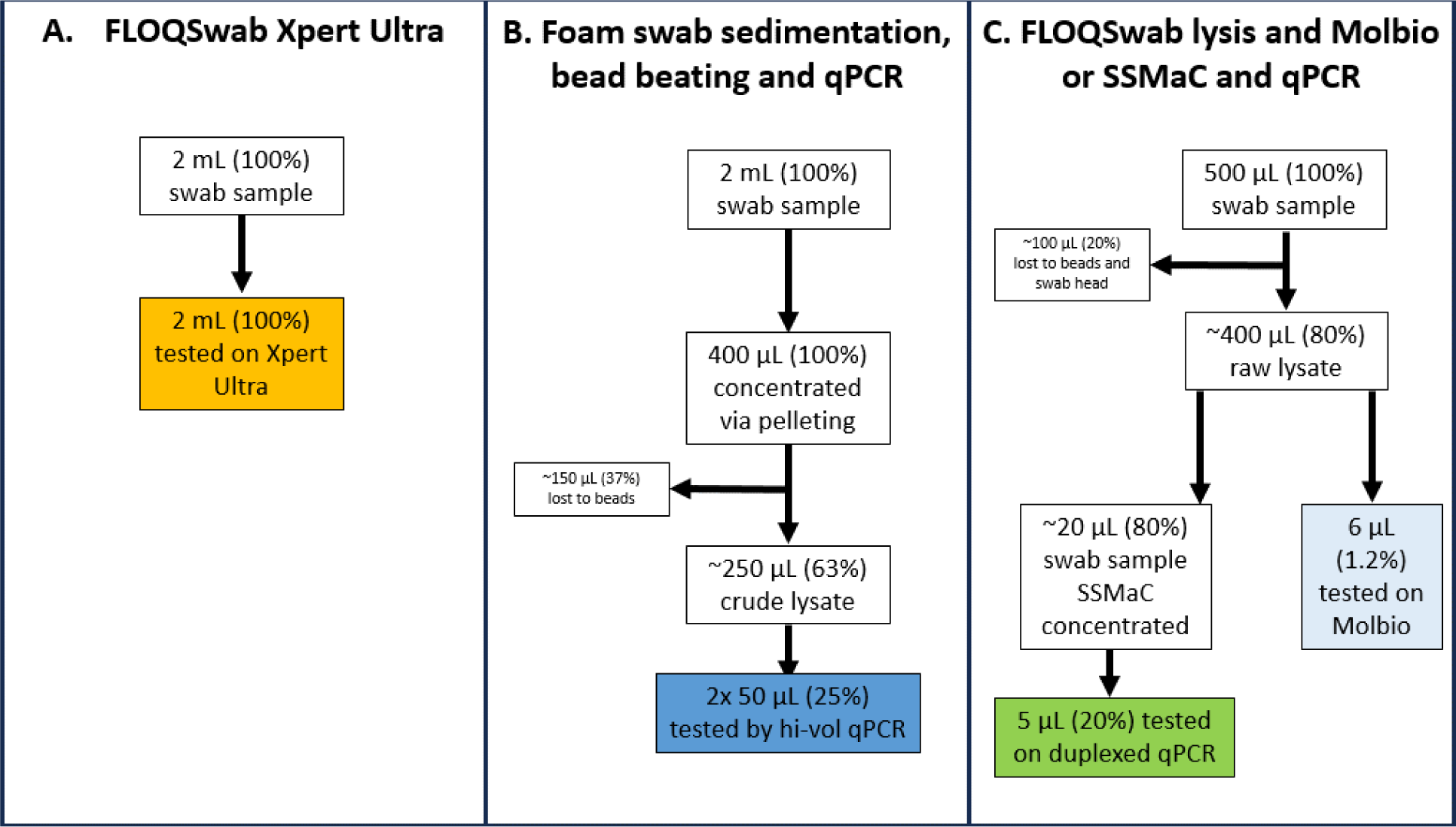
Sample volume to assay input volume flow chart

Prior to this analysis, it was not known whether the MTB DNA signal in oral swab samples was associated with bacterial cells, extracellular fractions, or both. When the concentrate and supernatant fractions were tested side by side by qPCR, the concentrated fraction had a significantly larger proportion of the detectable MTB DNA. Therefore, the majority of the TB signal observed in TS assays is associated with particulate material. These results do not allow us to definitively discern whether the MTB DNA resides within intact bacilli or is associated extracellularly with bacterial cells or other particulate material. Nevertheless, they indicate that simple sample concentration approaches such as centrifugation or filtration can be effective in tongue swab sample processing for TB detection.

This study had several limitations. The sample size was limited, and samples were processed in a non-blinded manner. The collection of foam swab samples was done after the collection of two FLOQSwabs. Although it is assumed that flocked swabs do not collect all available material from the tongue dorsum, we cannot know if the foam swabs would have performed differently if collected first. Samples were frozen for several months and shipped to a lab for processing; different results may have been observed if samples were tested fresh on site.

Despite these limitations, the results support strategies that may be useful for developing automated, POC technologies for TS. All the elements included in SSMaC are automatable and promising for developing a dedicated device. For example, the DASH system incorporates SSMaC and is designed specifically for testing swabs. Although centrifugation is challenging to automate, a POC filtration system like the one used in Xpert Ultra can achieve the same effect.

In conclusion, these results confirm and extend evidence from others (14) that TS testing can be highly sensitive and specific when purpose-designed methods are used to process and test the samples. With further development and automation, TS can become a valuable addition to the TB control toolkit, enabling high throughput, broadly tolerated sampling of virtually any individual in any setting. TS has the potential to simplify TB diagnosis, improve the care of many TB patients, and enable active case-finding strategies that will reduce TB transmission.

## Supporting information

Olson et al supplemental information

## Data Availability

Data have been uploaded to Mendeley Data, a cloud-based, generalist community data repository that is recommended by the NIH. Additional information is available upon reasonable request to the authors.

https://data.mendeley.com/

## Acknowledgements

This work was supported by the Bill & Melinda Gates Foundation (INV-004527, OPP 1213054), NIAID grants R01AI139254 and R21AI185407, and by NIBIB grant U54EB027049. Under the grant conditions of the Bill & Melinda Gates Foundation, a Creative Commons Attribution 4.0 Generic License has already been assigned to the Author Accepted Manuscript version that might arise from this submission. We thank Copan Italia for providing swabs.

