## Supplementary material for "High-sensitivity detection of *Mycobacterium tuberculosis* DNA in tongue swab samples": Olson et al supplemental information

**Supplemental material**

### Supplemental Material 1 (S1)

**Biomass proxy experiment.** Copan FLOQSwabs® Flocked Swabs **(**flocked swab) and Medline Pint-Size™ DenTips® Oral Swabs (foam swab) samples were collected from healthy volunteers in Seattle. Swabs were stored dry (without buffer) at -80 °C until ready for processing. Without thawing, frozen swabs were removed from the freezer and immediately boiled for 10 min at 100 °C. TE buffer was added as 0.5-mL applications. Swabs were vortexed on high for 30 sec, and the entire recoverable material was removed using a micropipette and transferred to a new 1.5-mL snap-cap tube. A NanoDrop® ND-1000 Spectrophotometer (version 3.8.1) was used to measure absorbency on the Nucleic Acid module. Results were exported to Microsoft Excel where they were further analyzed.

**Figure S1. Absorbance (LOG$_2$ $A_{260}$) average ± standard deviation from experimental foam and flocked swab tongue samples**

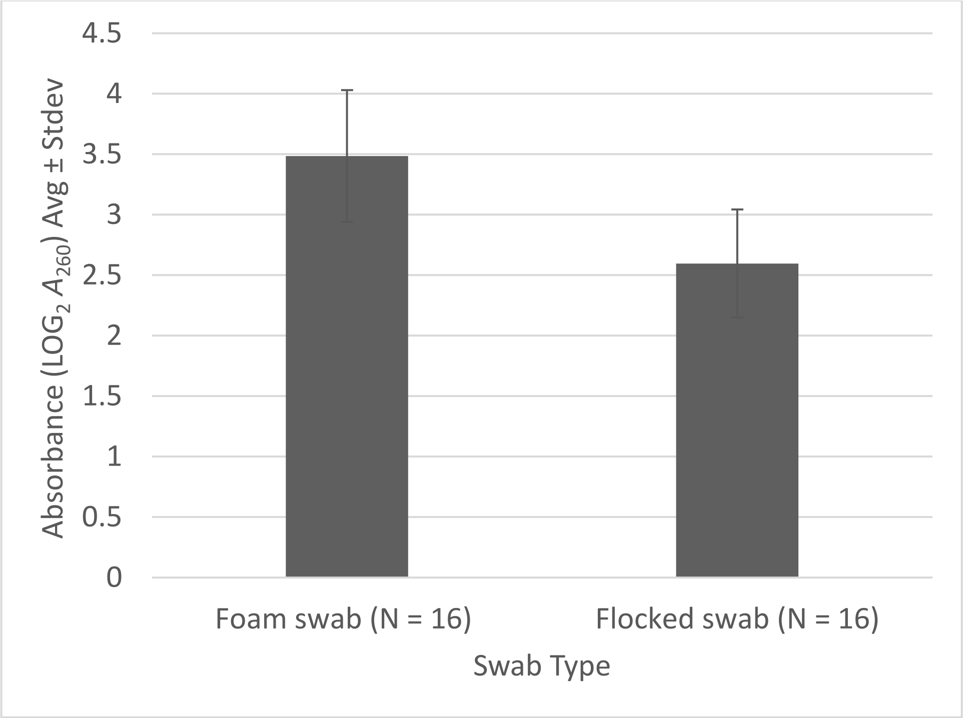


Swabs were collected from healthy donors in Seattle and boil prepped. Higher peaks at 260 nm (more nucleic acid) were observed in the foam samples compared to flocked swabs. A single-tailed, unpaired t-test was statistically significant ($p$ = 0.000016). Roughly twice as much nucleic acid is measured on average (3.49 ± 0.55) in the foam samples than the flocked swab samples (2.60 ± 0.45). Since the data is expressed exponentially a delta of 1 LOG$_2$ $A_{260}$ indicates a doubling between the two treatment groups; in this case we see a delta of 0.89. The two swab types do have slight standard deviation overlap. This experiment is limited by sample population. Yields may differ in swabs collected from people experiencing disease (TB or otherwise). Spectrophotometry is also a limited method to measure nucleic acid, especially in a minimally prepped sample type. As a proxy, nucleic acid from all of the oral biomass collected from the dorsum of the tongue could indicate which sample type would have more sample collected, and in theory, more TB cells (should they be present) in the final sample. Ultimately, the clinical foam samples performed well whether or not bacilli collection was doubled.

### Supplemental Material 2 (S2)

| **Name** | **Capture Probe Sequence** |
|---|---|
| 1081 FWD | **5’-**/52-Bio/AAAAAAAAAAAAAAAAAAAATACCGCCACCGTGATTTC/3SpC3/**-3’** |
| 1081 REV | **5’-**/52-Bio/AAAAAAAAAAAAAAAAAAAATCCGGGAAATAGCTGCC/3SpC3/**-3’** |
| 6110 FWD | **5’-**/52-Bio/AAAAAAAAAAAAAAAAAAAAGACCACCAGCACCTAACC/3SpC3/**-3’** |
| 6110 REV | **5’-**/52-Bio/AAAAAAAAAAAAAAAAAAAAGTGACAAAGGCCACGTAG/3SpC3/**-3’** |

/52-Bio/ - Dual biotin modification at 5’ end

/3SpC3/ - C3 spacer at 3’ end

AAAAAAAAAAAAAAAAAAAA– 20 nt poly(A) sequence at 5’ end
